# Use of robust norming to create a sensitive cognitive summary score in *de novo* Parkinson’s disease

**DOI:** 10.1101/2024.07.08.24310076

**Authors:** Daniel Weintraub, Michael C. Brumm, Ryan Kurth, Michele K. York, the Parkinson’s Progression Markers Initiative

## Abstract

**Background and Objectives:** Cognitive impairment is common at all stages in Parkinson’s disease (PD). However, the field is hampered by consensus over which neuropsychological tests to use and how to utilize the results generated by a cognitive battery. An option that combines the richness of a neuropsychological battery with the simplicity of a single test score is a cognitive summary score (CSS). The objective was to determine if a CSS created using robust norming is sensitive in detecting early cognitive deficits in *de novo*, untreated PD.

**Methods:** Using baseline cognitive data from PD participants and healthy controls (HCs) in the Parkinson’s Progression Markers Initiative, these steps were taken: (1) creating a robust HC subgroup that did not demonstrate cognitive decline over time; (2) using the robust HC subgroup to create regression-based internally-derived standardized scores (z-scores) for six cognitive scores across five tests; and (3) creating a CSS by averaging all standardized test z-scores.

**Results:** PD participants scored worse than HCs on all cognitive tests, with a larger effect size (PD versus HCs) when the comparison group was the robust HC subgroup compared with all HCs. Applying internally-derived norms rather than published norms, the largest cognitive domain effect sizes (PD vs. robust HCs) were for processing speed/working memory (Cohen’s d= -0.55) and verbal episodic memory (Cohen’s d= -0.48 and -0.52). In addition, using robust norming shifted PD performance from the middle of the average range (CSS z-score= -0.01) closer to low average (CSS z-score= -0.40), with the CSS having a larger effect size (PD vs. robust HC subgroup; Cohen’s d= -0.60) compared with all individual cognitive tests.

**Discussion:** PD patients perform worse cognitively than HC at disease diagnosis on multiple cognitive domains, particularly information processing speed and verbal memory. Using robust norming increases effect sizes and lowers the scores of PD patients to “expected” levels. The CSS performed better than all individual cognitive tests. A CSS developed using a robust norming process may be sensitive to cognitive changes in the earliest stages of PD and have utility as an outcome measure in clinical research, including clinical trials.

## INTRODUCTION

Long-term significant cognitive impairment is a common and dreaded outcome in Parkinson’s disease (PD), with dementia affecting up to 80% of patients long-term^1,2^. Mild cognitive impairment (MCI) is also relatively common^3^, even in *de novo* or early disease^4-6^. In addition, subtle cognitive changes can be detected in the prodromal state, including in patients with isolated REM sleep behavior disorder (iRBD)^7^ or persons with hyposmia^8^.

However, the ability to track cognitive changes over time, including in the context of short-term randomized controlled trials (RCTs), is hampered by consensus over which neuropsychological tests to use and how to utilize the results^9,10^. This may have contributed to having only one cognitive enhancing medication FDA-approved for the treatment of PD dementia (rivastigmine)^11^ with no other positive RCTs in the 20 years since then, and no positive RCTs for PD-MCI^12^.

Increasingly computerized cognitive batteries and smartphone-based cognitive apps are being used in PD, but most are not yet fully validated and ready for routine use in clinical research. Global screening instruments commonly used in PD (e.g., Mini-Mental State Examination [MMSE]^13^ and Montreal Cognitive Assessment [MoCA])^14^ are brief (i.e., 5-10 minutes), but may not be sensitive to mild cognitive changes in the short-term or may have significant variability.

Many individual paper-and-pencil cognitive tests are used in PD, both in clinical care and clinical research, and they are commonly assembled into a cognitive battery. Given the heterogeneity of cognitive deficits in PD, even at the stage of MCI^3^, batteries often assess multiple cognitive domains. Many versions of detailed cognitive batteries (e.g., recommended Level I or Level II PD-MCI batteries^9^) are utilized, with some evidence that there is differential sensitivity among commonly-used neuropsychological tests to mild cognitive deficits in PD^15^. Recent research has suggested that in general Level I and Level II cognitive batteries are better able to predict conversion from PD-MCI to PD dementia than are global screening instruments^16^. However, it remains unclear how to best utilize and interpret results containing multiple individual (sub)scores from differently-scaled and - normed tests.

An option that allows one to combine the richness and details of a neuropsychological battery with the simplicity of a single test score is to create a cognitive summary score (CSS) after all detailed test results are normed on the same scale (e.g., z-score). Examples from other neurodegenerative diseases include: (1) Alzheimer’s disease (AD): the Preclinical Alzheimer Cognitive Composite (PACC; consisting of total recall score from the Free and Cued Selective Reminding Test, delayed recall score on the Logical Memory IIa sub-test from the Wechsler Memory Scale, Digit Symbol Substitution Test and MMSE total score)^17^, and the AD Composite Score (ADCOMS; 4 Alzheimer’s Disease Assessment Scale–cognitive subscale items, 2 Mini-Mental State Examination items, and all 6 Clinical Dementia Rating—Sum of Boxes items)^18^; and (2) Huntington’s disease (HD): the Huntington’s Disease Cognitive Assessment Battery (HD-CAB; consisting of Symbol Digit Modalities Test, Paced Tapping, One Touch Stockings of Cambridge (abbreviated), Emotion Recognition, Trail Making B and Hopkins Verbal Learning Test)^19^.

In the Parkinson Associated Risk Syndrome (PARS) study a CSS was generated from a very detailed cognitive battery and used to document subtle cognitive changes in prodromal PD (i.e., persons with hyposmia + dopamine transporter [DAT] SPECT scan deficit)^8,20^. To our knowledge, there have not been other published literature documenting attempts to create a cognitive composite from detailed individual tests.

If determined to be more sensitive to early cognitive decline in PD, it is possible that a CSS would allow more neurobiological predictors of cognitive decline to be identified, and even serve as an outcome measure for RCTs, including in persons with prodromal or at-risk PD. In addition, such a CSS could be valuable in the routine, clinical evaluation of PD patients, as clinical neuropsychological evaluations often administer the battery of standard neuropsychological tests utilized in PPMI.

The Parkinson’s Progression Markers Initiative (PPMI) study originally had a PD-MCI Level I battery (five detailed, well-established cognitive tests) with published norms^21^. Given that PPMI PD participants are highly educated and motivated, we implemented robust norming to increase sensitivity to detect cognitive differences in PD compared with healthy controls (HCs), in a fashion similar to that employed to update the norms for the Dementia Rating Scale^22^. We hypothesized that using robust norming would increase sensitivity and adjust PD participant scores to the expected slight impairment reported in the literature^6^, and that a cognitive summary score would be more sensitive than individual test results in detecting cognitive differences in *de novo*, untreated PD patients compared with HCs.

## METHODS

### Standard protocol approvals, registrations and patient consents

An ethical standards committee on human experimentation reviewed and approved the study at each site. Written informed consent was obtained from all participants in the study.

### Cohort

The PPMI study and cohort has been extensively described^23,24^. All participant signed an approved informed consent form. Inclusion criteria for PD participants included: (1) an asymmetric resting tremor or asymmetric bradykinesia, or at least two symptoms out of resting tremor, bradykinesia, and rigidity; (2) a recent clinical diagnosis of PD (mean [SD] duration from diagnosis = 8.5 [7.3] months); (3) being untreated with PD medications; (4) evidence of dopaminergic deficit based on DAT SPECT imaging; and (5) being non-demented. HCs were required to (1) not have clinically significant neurological dysfunction, (2) not have a first-degree relative with PD and (3) have a Montreal Cognitive Assessment (MoCA) score ≥27. Participants enrolled as HCs who had evidence of CSF α-synuclein seed amplification assay (SAA) positivity were not considered to be HCs for the purposes of this analysis.

### Creating database

Baseline data were used, at which point PD participants were recently diagnosed and had not yet begun treatment with PD medication. The original cognitive battery was used to create the CSS to optimize the amount of data available for analyses. This original battery was composed of the Hopkins Verbal Learning Test-Revised (HLVT-R immediate and delayed free recall scores)(24), the Benton Judgment of Line Orientation -15 item version (JLO)(25), Symbol-Digit Modalities Test (SDMT)(26), Letter-Number Sequencing (LNS)(27), and semantic (animal) fluency(28). Together these tests assess memory, visuospatial function, information processing speed, executive function, working memory and language.

### Published norms

Using previously published methods derived from participants not enrolled in PPMI, “external” standard scores were derived. This included T-scores for the HVLT-R (immediate and delayed free recall scores), SDMT and semantic fluency, and scaled scores for the JLO and LNS.

### Robust norming

The robust HC group was defined using the criteria depicted in **Figure 1**. Briefly, participants were required to have a baseline MoCA of ≥27, have a year 1 MoCA score of ≥26, and not have had more than a 2-point drop in their MoCA score between baseline and year 1. This subset of participants was used to create internal norms for each test which were then used to calculate internally-derived z-scores for each participant (see Statistical Analysis for details).

**Figure 1.**
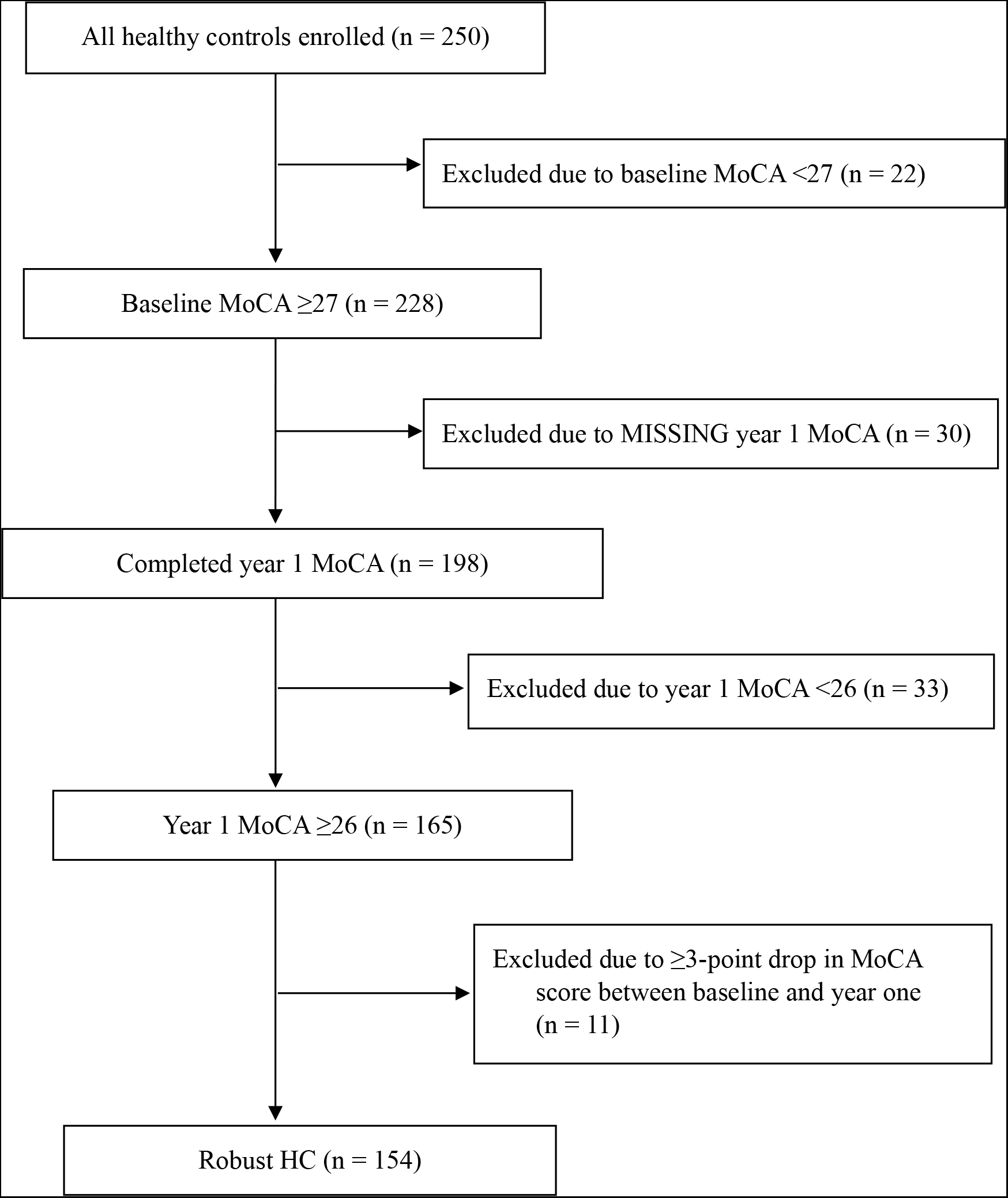
CONSORT Diagram for Super Healthy Controls.

### Data Availability

PPMI data used in the preparation of this article were obtained November 6, 2023 from the Parkinson’s Progression Markers Initiative (PPMI) database (www.ppmi-info.org/access-data-specimens/download-data), RRID:SCR 006431. This analysis was conducted by the PPMI Statistics Core and used actual dates of activity for participants, a restricted data element not available to public users of PPMI data. PPMI data are publicly available from the Parkinson’s Progression Markers Initiative (PPMI) database (www.ppmi-info.org/access-data-specimens/download-data). For up-to-date information on the study, visit www.ppmi-info.org.

### Statistical Analysis

Statistical analyses were performed using SAS v9.4 (SAS Institute Inc., Cary, NC; sas.com; RRID:SCR_008567). Statistical analysis codes used to perform the analyses in this article are shared on Zenodo [10.5281/zenodo.12636045].

All external standard scores derived from published norms (T-scores or scaled scores) were converted to z-scores and then capped at -3 to 3. To derive internal norms, the robust HC population was used to build a linear regression model for each test. Age, sex, and continuous education were considered as possible predictors of raw test scores. The second order polynomials of centered age and education were also considered. Sex was coded as a binomial variable with male = 1 and female = 2. Backwards selection with a cutoff of alpha = 0.10 was used to determine which variables would appear in each final model. The Kolmogorov–Smirnov test was used to assess the normality of the residuals of each model. These derived linear models were used to calculate an “expected” raw score for each participant and each test, and then a z-score was calculated by taking the participant’s raw score minus their expected score and dividing by the root mean square error. These internal norms were also capped at -3 to 3.

Two cognitive summary scores were calculated for each participant, one being an average of their six external z-scores (derived from published norms), and the other an average of their six internal z-scores (derived from the PPMI robust HC subgroup), with the six z-scores weighted equally in both cases.

For each variable in Table 1, two-sample t-tests and chi-squared tests were performed to compare the PD group to each of the HC groups. Cohen’s d effect sizes were used to compare these composite scores across groups.

**Table 1.**
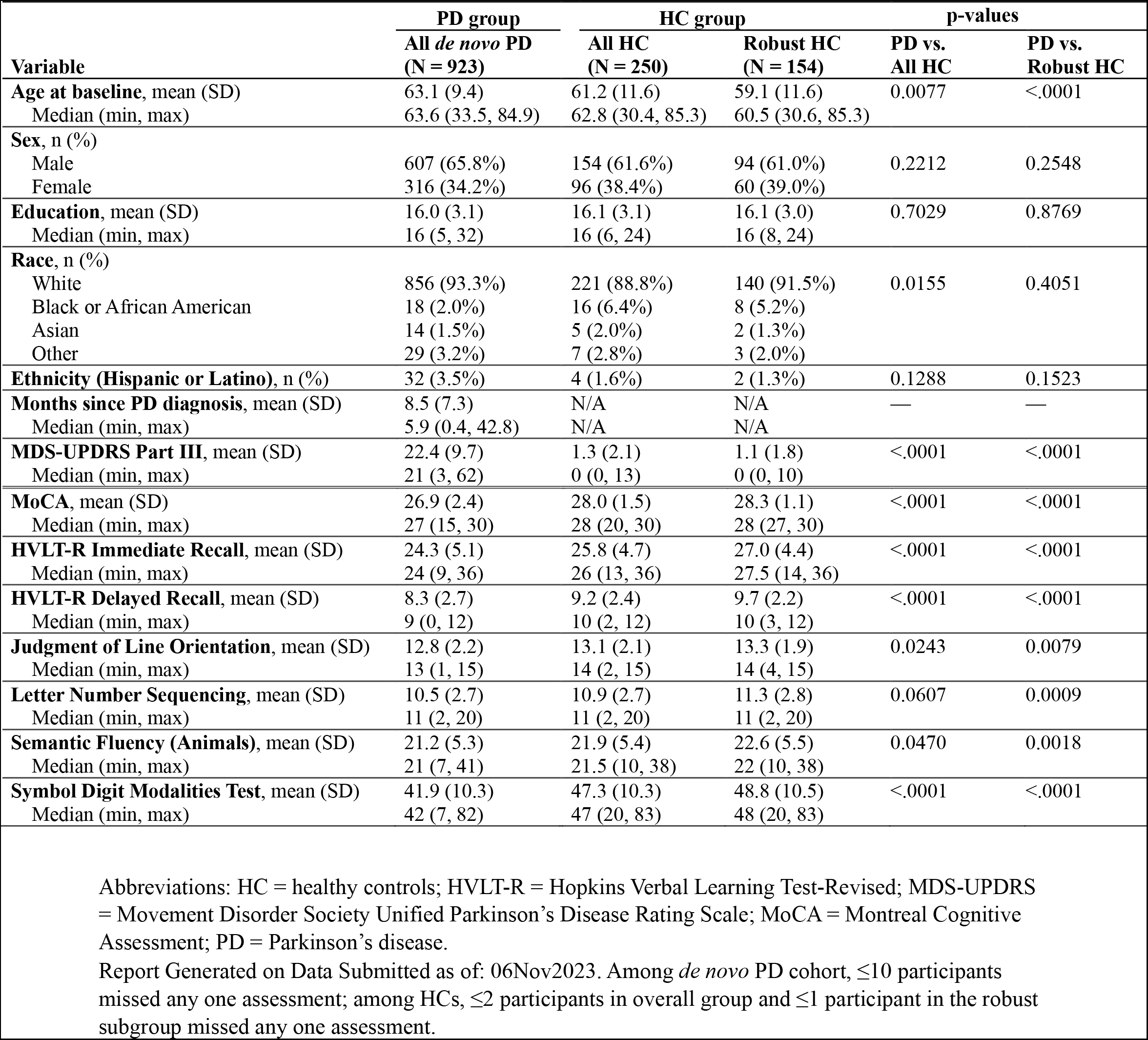
Demographic characteristics of PD and HC cohorts.

## RESULTS

### Cohort characteristics

There were 923 participants with *de novo*, untreated PD; they had mean disease duration <1 year, mean age of 63 years, and were two-thirds male, highly educated, and overwhelmingly white and non-Hispanic (**Table 1**). There were 250 total HCs, and 154 participants in the robust HC subgroup. PD participants were older than the total HCs (mean age = 61) and robust HCs (mean age = 59), but otherwise had similar demographic characteristics.

### Raw cognitive test scores

PD participants scored worse on global cognition (MoCA score) by approximately 1 point compared with both HC groups, as well as worse on all detailed cognitive test raw scores, except LNS for the entire HC group (**Table 1**). The entire HC group scored worse numerically on all cognitive tests compared with the robust HC subgroup.

### Using robust HC subgroup instead of entire HC group

PD participants scored worse than the entire HC group and the robust HC subgroup on all detailed cognitive tests using both published norms (**Table 2**) and internally-derived norms (**Table 3**). The effect size (Cohen’s d) was greater (sometimes twice as great) when comparing the PD group to the robust HC subgroup rather than the entire HC group for all cognitive tests, whether using published norms (**Table 2**) or internally-derived norms (**Table 3**). The tests with the biggest effect sizes, which were medium-sized, were for information processing speed (SDMT) and verbal episodic learning and memory (HVLT-R). Smaller effect sizes were seen for JLO, LNS and semantic fluency, but the effect size was increased for these tests by using the robust HC subgroup as the comparator group.

**Table 2.**
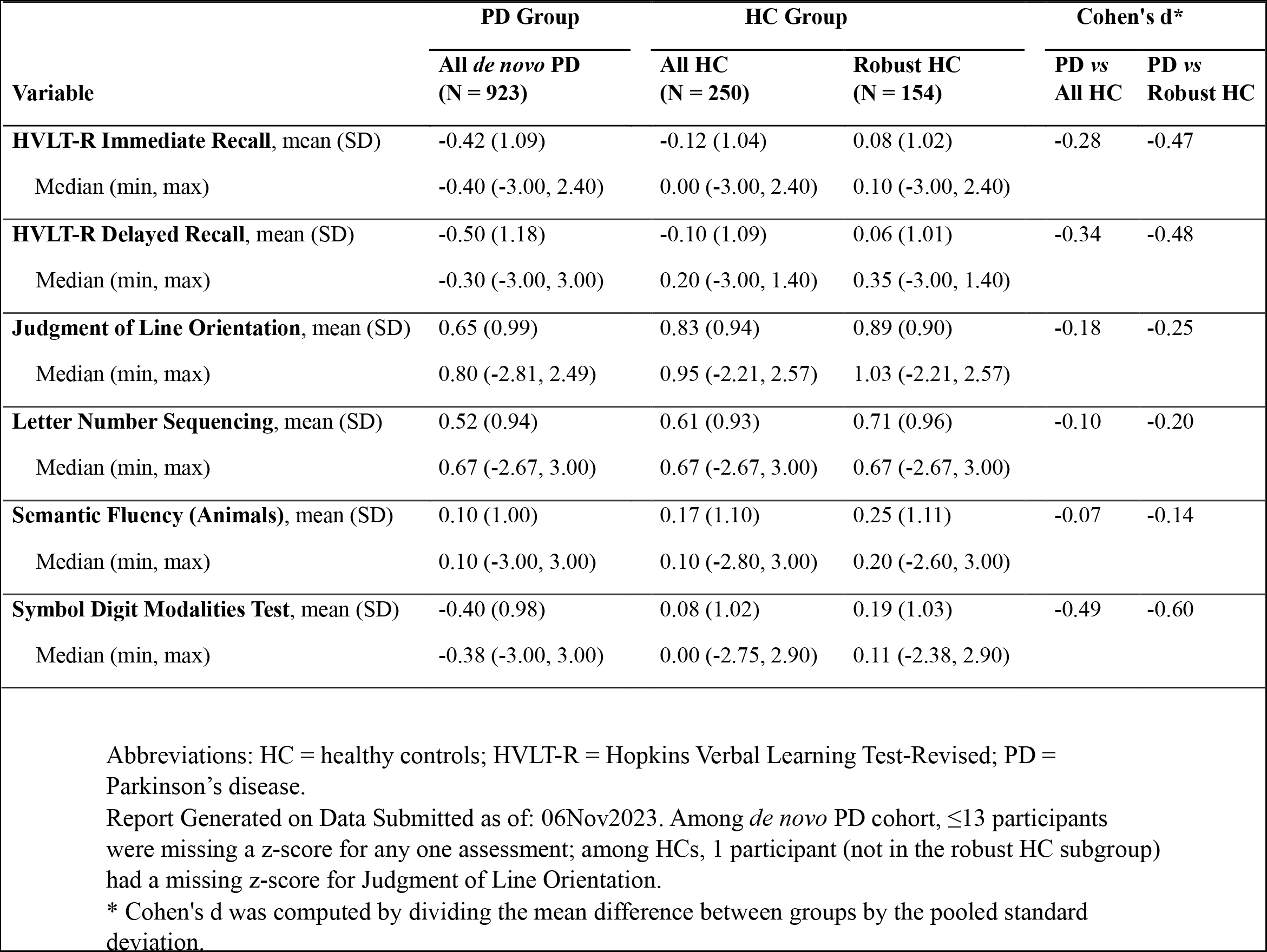
Published norms z-scores at baseline among de novo PD and HC cohorts.

**Table 3.**
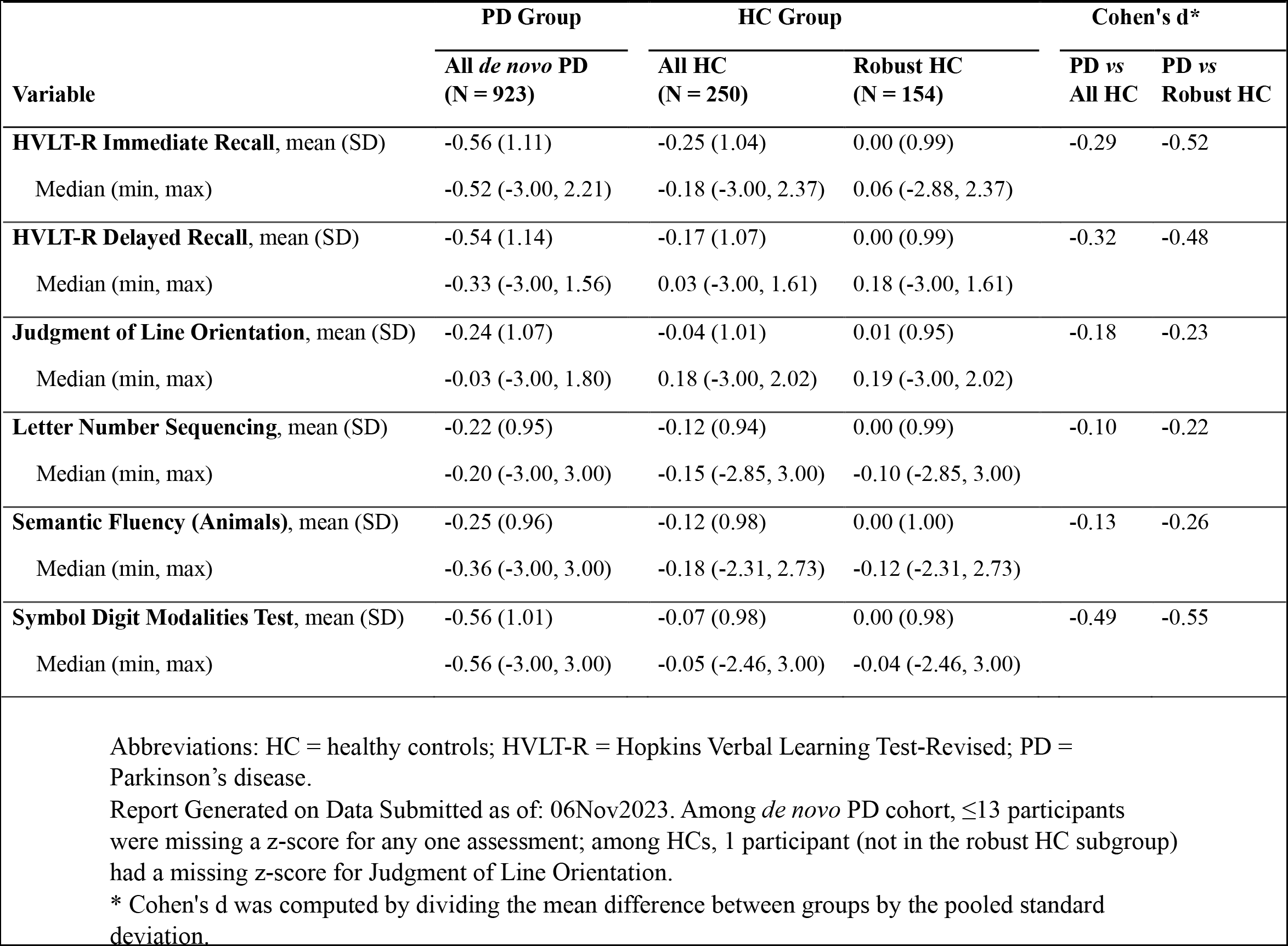
Internally-derived, regression-based z-scores at baseline among de novo PD and HC cohorts.

### Regression-based norms derived from robust healthy controls

The results of regression analyses of the effects of age, sex, and education on the raw test scores of robust SH participants are summarized in **Supplementary Table 1**. In brief, significant predictors included age and sex for the immediate and delayed recall scores of the HVLT-R; age, sex, and education for the JLO; age and education for the LNS; only sex only for semantic fluency; and only age for the SDMT. For all tests where age was a significant predictor, older participants scored lower on average. Females scored significantly higher on the HVLT-R and semantic fluency; whereas males scored higher on the JLO. Participants with higher education performed better on the JLO and LNS. Kolmogorov–Smirnov tests (results not shown) indicated normally distributed residuals for HVLT-R immediate recall, LNS, SDMT, and semantic fluency; however, residuals were left-skewed for HVLT-R delayed recall (skewness = -0.80; D = 0.09; *p* < 0.01) and JLO (skewness = -1.36; D = 0.11; *p* < 0.01).

### Using internally-derived norms instead of published norms

The effect sizes for PD versus both all HC and the robust HC subgroup were similar for internally-derived (**Table 3**) vs. published norms (**Table 2**), except for semantic fluency, for which the effect size was approximately twice as great when using internally-generated norms. The main change occurring with internally-derived norms was a shift for nearly all tests (except HVLT-R delayed recall, which was relatively unchanged) to a more negative (i.e., more impaired) z-score, for both PD participants and HCs. Specifically, z-scores for PD participants ranged from -0.50 to +0.65 using published norms, but -0.56 to -0.22 using internally-derived norms.

### Cognitive summary score

Using published norms, the CSS showed a larger effect size than all individual tests for both HC groups, except for the SDMT (**Table 4**). Using internally-derived norms, the CSS had a larger effect size than all individual tests when using the robust HC subgroup as the comparator group (**Table 4**). Once again, changing from published to internally-derived norms decreased the z-score for the CSS from average (−0.01) to closer to low average (−0.40).

**Table 4.** Cognitive summary score using published and internally-derived norms.

| Variable | PD Group | HC Group |  | Cohen's d* |  |
| --- | --- | --- | --- | --- | --- |
|  | All *de novo* PD<br>(N = 923) | All HC<br>(N = 250) | Robust HC<br>(N = 154) | PD vs<br>All HC | PD vs<br>Robust HC |
| **Published norms** |  |  |  |  |  |
| CSS, mean (SD) | -0.01 (0.64) | 0.25 (0.61) | 0.36 (0.61) | -0.42 | -0.59 |
| Median (min, max) | 0.02 (-2.34, 1.99) | 0.26 (-1.73, 1.99) | 0.32 (-1.73, 1.99) |  |  |
| **Internally-derived norms** |  |  |  |  |  |
| CSS, mean (SD) | -0.40 (0.68) | -0.12 (0.62) | 0.00 (0.61) | -0.41 | -0.60 |
| Median (min, max) | -0.34 (-2.75, 1.76) | -0.12 (-1.85, 1.83) | -0.03 (-1.85, 1.83) |  |  |

Abbreviations: CSS = cognitive summary score; HC = healthy controls; PD = Parkinson's disease.
Report Generated on Data Submitted as of: 06Nov2023. Among *de novo* PD cohort, 17 participants had a missing value for both cognitive summary scores; among HCs, 1 participant (not in the robust HC subgroup) had a missing value for both scores.
\* Cohen's d was computed by dividing the mean difference between groups by the pooled standard deviation.

## DISCUSSION

Using PPMI baseline cognitive data from *de novo*, untreated PD patients and demographically-similar HCs, we demonstrated that it is possible to create a CSS using robust norming that is sensitive to subtle cognitive changes in early PD.

Robust norming has been used to update the norms of commonly-used cognitive tests^25^, given that studies have indicated that cross-sectional normative samples of older adults include a mix of cognitively normal individual but also those in the initial stages of cognitive decline. This contaminates cross-sectional normative samples with cases of preclinical dementia that leads to an underestimation of the test mean and overestimation of the variance, thus reducing their clinical utility. When the DRS was renormed, use of the robust norms resulted in an almost twofold increase in sensitivity with relatively minimal loss of specificity when compared with the original norms^22^.

Cognitive summary scores, also call cognitive composites, have been developed and utilized in clinical research for both AD and HD, including RCTs. The ADCOMS, developed to be sensitive to the early stages of AD and to changes with symptomatic therapy^18^, showed statistically significant improvement with lecanemab treatment in a recent AD trial^26^. The PACC has been shown to be sensitive to beta amyloid (Aβ) positivity in cognitively-unimpaired individuals^27^, although it was negative as the primary outcome in a trial of solanezumab for preclinical AD^28^. For HD, the HD-CAB, which yields a composite z-score that is sensitive in pre-HD and early HD group, and shows high test-retest reliability^19^, is recommended for use in clinical care and research, but may need modifications and there aren’t published results reporting on its use in a RCT^29^.

We found that PD patients performed worse than HCs, whether using raw scores, published norms or internally-derived norms. This is consistent with what has been reported in other studies of newly-diagnosed PD cohorts^5^, and with research showing that 15-30% of *de novo* patients meet criteria for MCI^6^.

PD patients also performed worse cognitively than HCs on all cognitive domains assessed, which is also consistent with multiple domains being impaired at the stage of MCI in PD^3^, which makes the case that any cognitive battery in PD should assess multiple cognitive domains. Regarding specific cognitive tests, the most notable differences when comparing PD patients with HCs was for SDMT (information processing speed) and HVLT-R (verbal learning and memory), suggesting that these tests are good candidates for inclusion in a PD cognitive battery.

As part of robust norming, we excluded HCs who experienced cognitive decline over the first year of the study or had evidence for neuronal alpha-synuclein disease based on their CSF alpha-synuclein seed amplification assay test. Using the robust HC subgroup instead of the entire HC group significantly increased the effect sizes of cognitive differences between PD patients and HCs, confirming the value in excluding HCs with incipient cognitive decline from the norming process.

The second stage of the robust norming process was the creation of internally-derived norms instead of relying on published norms obtained from different populations, which is important given that the PPMI cohort, both PD patients and HCs, are highly educated (82% of the PD patients reported having formal education beyond high school, and only two cognitive tests in the battery adjust for education), motivated and younger than some other newly-diagnosed cohorts. Using the internally-derived versus published norms lowered the standardized test scores of PD patients to “expected” levels (between average and low average), and allowed direct comparison across the tests due to the same reference population being used, as opposed to different reference populations for most published norms.

The final step after robust norming was to create a CSS by averaging the individual test z-scores generated by robust norming. When doing this, and using the robust HC subgroup as the comparator, the cognitive summary score had a greater effect size than any of the individual tests comprising the battery, showing enhanced sensitivity for a summary score when using a robust HC group. This process also allows for applying z-score cut-offs to define cognitive impairment, which might be used as an inclusion criterion or as an endpoint in clinical trials.

Future research for a cognitive summary score in PD might include using other cognitive tests to compare performance against this battery, testing it against a global screening instrument (e.g., the MoCA), considering unequal weighting of individual test scores based on their effect size, testing it in prodromal PD (e.g., iRBD or hyposmia), determining how the cognitive summary score evolves and predicts outcomes over time, determining its association with biological variables and using it as an outcome measure in clinical trials.

Strengths of the study include a very large sample size, a multi-site international cohort, and use of well-validated and commonly-used neuropsychological tests. Limitations include not having a Level II PD-MCI cognitive battery and having different inclusion criteria at baseline for PD patients and HCs (i.e., HCs had to have MoCA score ≥27, while PD patients did not).

In conclusion, robust norming enhanced sensitivity in detecting cognitive differences in *de novo*, untreated PD, and generated scores consistent with the mild cognitive deficits known to commonly occur in prodromal and early disease. The CSS generated after the robust norming process combines the richness of a detailed, multi-domain cognitive battery with the simplicity a single, identically-normed and equally-weighted score with potential for domain subscores.

## Supporting information

PPMI Study Group Authors

## Acknowledgments

PPMI – a public-private partnership – is funded by the Michael J. Fox Foundation for Parkinson’s Research and funding partners, including 4D Pharma, Abbvie, AcureX, Allergan, Amathus Therapeutics, Aligning Science Across Parkinson’s, AskBio, Avid Radiopharmaceuticals, BIAL, BioArctic, Biogen, Biohaven, BioLegend, BlueRock Therapeutics, Bristol-Myers Squibb, Calico Labs, Capsida Biotherapeutics, Celgene, Cerevel Therapeutics, Coave Therapeutics, DaCapo Brainscience, Denali, Edmond J. Safra Foundation, Eli Lilly, Gain Therapeutics, GE HealthCare, Genentech, GSK, Golub Capital, Handl Therapeutics, Insitro, Jazz Pharmaceuticals, Johnson & Johnson Innovative Medicine, Lundbeck, Merck, Meso Scale Discovery, Mission Therapeutics, Neurocrine Biosciences, Neuron23, Neuropore, Pfizer, Piramal, Prevail Therapeutics, Roche, Sanofi, Servier, Sun Pharma Advanced Research Company, Takeda, Teva, UCB, Vanqua Bio, Verily, Voyager Therapeutics, the Weston Family Foundation and Yumanity Therapeutics.

---

**Supplementary Table 1.** Regression models among robust healthy controls.

| Test | Variables | *B* | SE | *t* | *p* | Adj R<sup>2</sup> | RMSE |
| --- | --- | --- | --- | --- | --- | --- | --- |
| **HVLT-R Immediate Recall** | Intercept | 28.37542 | 2.157 | 13.154 | <.001 | 0.047 | 4.32146 |
|  | Age | -0.05979 | 0.030 | -1.972 | 0.050 |  |  |
|  | Sex* | 1.58306 | 0.717 | 2.208 | 0.029 |  |  |
| **HVLT-R Delayed Recall** | Intercept | 10.92487 | 1.060 | 10.309 | <.001 | 0.070 | 2.12293 |
|  | Age | -0.03996 | 0.015 | -2.682 | 0.008 |  |  |
|  | Sex* | 0.80139 | 0.352 | 2.275 | 0.024 |  |  |
| **Judgment of Line Orientation** | Intercept | 15.07467 | 1.184 | 12.731 | <.001 | 0.132 | 1.77892 |
|  | Age | -0.02374 | 0.012 | -1.899 | 0.059 |  |  |
|  | Sex* | -1.29422 | 0.297 | -4.354 | <.001 |  |  |
|  | Education | 0.08678 | 0.048 | 1.800 | 0.074 |  |  |
| **Letter Number Sequencing** | Intercept | 12.30761 | 1.572 | 7.830 | <.001 | 0.056 | 2.68988 |
|  | Age | -0.05359 | 0.019 | -2.845 | 0.005 |  |  |
|  | Education | 0.13650 | 0.072 | 1.886 | 0.061 |  |  |
| **Semantic Fluency (Animal)** | Intercept | 20.25284 | 1.338 | 15.142 | <.001 | 0.016 | 5.49634 |
|  | Sex* | 1.71525 | 0.908 | 1.889 | 0.061 |  |  |
| **Symbol Digit Modalities Test** | Intercept | 76.80621 | 3.772 | 20.360 | <.001 | 0.268 | 8.95934 |
|  | Age | -0.47289 | 0.063 | -7.554 | <.001 |  |  |

Abbreviations: HC = healthy controls; HVLT-R = Hopkins Verbal Learning Test-Revised; PD = Parkinson's disease; RMSE = root mean square error.
Report Generated on Data Submitted as of: 06Nov2023.
\* Defined as 1=male and 2=female.

