## Supplementary material for "Use of robust norming to create a sensitive cognitive summary score in *de novo* Parkinson’s disease": PPMI Study Group Authors

### PPMI STUDY TEAMS/CORES/COLLABORATORS FOR PUBLICATIONS

**Executive Steering Committee:**
Kenneth Marek, MD[1] (Principal Investigator); Caroline Tanner, MD, PhD[9]; Tanya Simuni, MD[3]; Andrew Siderowf, MD, MSCE[12]; Douglas Galasko, MD[27]; Lana Chahine, MD[39]; Christopher Coffey, PhD[4]; Kalpana Merchant, PhD[59]; Kathleen Poston, MD[38]; Roseanne Dobkin, PhD[41]; Tatiana Foroud, PhD[15]; Brit Mollenhauer, MD[8]; Dan Weintraub, MD[12]; Ethan Brown, MD[9]; Karl Kieburtz, MD, MPH[23]; Mark Frasier, PhD[6]; Todd Sherer, PhD[6]; Sohini Chowdhury, MA[6]; Roy Alcalay, MD[35] and Aleksandar Videnovic, MD[45]

**Steering Committee:**
Duygu Tosun-Turgut, PhD[9]; Werner Poewe, MD[7]; Susan Bressman, MD[14]; Jan Hammer[15]; Raymond James, RN[22]; Ekemini Riley, PhD[40]; John Seibyl, MD[1]; Leslie Shaw, PhD[12]; David Standaert, MD, PhD[18]; Sneha Mantri, MD, MS[60]; Nabila Dahodwala, MD[12]; Michael Schwarzschild[45]; Connie Marras[43]; Hubert Fernandez, MD[25]; Ira Shoulson, MD[23]; Helen Rowbotham[2]; Paola Casalin[11] and Claudia Trenkwalder, MD[8]

**Michael J. Fox Foundation (Sponsor):** Todd Sherer, PhD; Sohini Chowdhury, MA; Mark Frasier, PhD; Jamie Eberling, PhD; Katie Kopil, PhD; Alyssa O'Grady; Maggie McGuire Kuhl; Leslie Kirsch, EdD and Tawny Willson, MBS

**Study Cores, Committees and Related Studies:** ***(Include as applicable to the paper)***
*Project Management Core:* Emily Flagg, BA[1]
*Site Management Core:* Tanya Simuni, MD[3]; Bridget McMahon, BS[1]
Strategy and Technical Operations: Craig Stanley, PhD[1]; Kim Fabrizio, BA[1]
*Data Management Core:* Dixie Ecklund, MBA, MSN[4]; Trevis Huff, BSE[4]
*Screening Core:* Tatiana Foroud, PhD[15]; Laura Heathers, BA[15]; Christopher Hobbick, BSCE[15]; Gena Antonopoulos, BSN[15]
*Imaging Core:* John Seibyl, MD[1]; Kathleen Poston, MD[38]
*Statistics Core*: Christopher Coffey, PhD[4]; Chelsea Caspell-Garcia, MS[4]; Michael Brumm, MS[4]
*Bioinformatics Core*: Arthur Toga, PhD[10]; Karen Crawford, MLIS[10]
*Biorepository Core:* Tatiana Foroud, PhD[15]; Jan Hamer, BS[15]
*Biologics Review Committee*: Brit Mollenhauer[8]; Doug Galasko[27]; Kalpana Merchant[59]
*Genetics Core:* Andrew Singleton, PhD[13]
*Pathology Core:* Tatiana Foroud, PhD[15]; Thomas Montine, MD, PhD[38]
*Found:* Caroline Tanner, MD PhD[9]
*PPMI Online:* Carlie Tanner, MD PhD[9]; Ethan Brown, MD[9]; Lana Chahine, MD[39]; Roseann Dobkin, PhD[41]; Monica Korell, MPH[9]

**Site Investigators:**

Charles Adler, PhD[49]; Roy Alcalay, MD[35]; Amy Amara, PhD[50]; Paolo Barone, PhD[30]; Bastiaan Bloem, PhD[58] Susan Bressman, MD[14]; Kathrin Brockmann, MD[26]; Norbert Brüggemann, MD[57]; Lana Chahine, MD[39]; Kelvin Chou, MD[42]; Nabila Dahodwala, MD[12]; Alberto Espay, MD[32]; Stewart Factor, DO[16]; Hubert Fernandez, MD[25]; Michelle Fullard, MD[50]; Douglas Galasko, MD[27]; Robert Hauser, MD[19]; Penelope Hogarth, MD[17]; Shu-Ching Hu, PhD[21]; Michele Hu, PhD[56]; Stuart Isaacson, MD[31]; Christine Klein, MD[57]; Rejko Krueger, MD[2]; Mark Lew, MD[47]; Zoltan Mari, MD[54]; Connie Marras, PhD[43]; Maria Jose Martí, PhD[33]; Nikolaus McFarland, PhD[52]; Tiago Mestre, PhD[44]; Brit Mollenhauer, MD[8]; Emile Moukheiber, MD[28]; Alastair Noyce, PhD[61] Wolfgang Oertel, PhD[62]; Njideka Okubadejo, MD[63]; Sarah O'Shea, MD[37]; Rajesh Pahwa, MD[46]; Nicola Pavese, PhD[55]; Werner Poewe, MD[7]; Ron Postuma, MD[53]; Giulietta Riboldi, MD[51]; Lauren Ruffrage, MS[18]; Javier Ruiz Martinez, PhD[34]; David Russell, PhD[1]; Marie H Saint-Hilaire, MD[22]; Neil Santos, BS[49]; Wesley Schlett[45]; Ruth Schneider, MD[23]; Holly Shill, MD[48]; David Shprecher, DO[24]; Tanya Simuni, MD[3]; David Standaert, PhD[18]; Leonidas Stefanis, PhD[36]; Yen Tai, PhD[29]; Caroline Tanner, PhD[9]; Arjun Tarakad, MD[20]; Eduardo Tolosa PhD[33] and Aleksandar Videnovic, MD[45]

**Coordinators:**

Susan Ainscough, BA[30]; Courtney Blair, MA[18]; Erica Botting[19]; Isabella Chung, BS[54]; Kelly Clark[24]; Ioana Croitoru[34]; Kelly DeLano, MS[32]; Iris Egner, PhD[7]; Fahrial Esha, BS[51]; May Eshel, MSc[35]; Frank Ferrari, BS[42]; Victoria Kate Foster[55]; Alicia Garrido, MD[33]; Madita Grümmer[57]; Bethzaida Herrera[48]; Ella Hilt[26]; Chloe Huntzinger, BA[50]; Raymond James, BS[22]; Farah Kausar, PhD[9]; Christos Koros, MD, PhD[36]; Yara Krasowski, MSc[58]; Dustin Le, BS[17]; Ying Liu, MD[50]; Taina M. Marques, PhD[2]; Helen Mejia Santana, MA[37]; Sherri Mosovsky, MPH[39]; Jennifer Mule, BS[25]; Philip Ng, BS[43]; Lauren O'Brien[46]; Abiola Ogunleye, PGDip[29]; Oluwadamilola Ojo, MD[63]; Obi Onyinanya, BS[28]; Lisbeth Pennente, BA[31]; Romina Perrotti[53]; Michael Pileggi, MS[53]; Ashwini Ramachandran, MSc[12]; Deborah Raymond, MS[14]; Jamil Razzaque, MS[56]; Shawna Reddie, BA[44]; Kori Ribb, BSN,[28]; Kyle Rizer, BA[52]; Janelle Rodriguez, BS[27]; Stephanie Roman, HS[1]; Clarissa Sanchez, MPH[20]; Cristina Simonet, PhD[29]; Anisha Singh, BS[23]; Elisabeth Sittig, RN[62]; Barbara Sommerfeld MSN[16]; Angela Stovall, BS[42]; Bobbie Stubbeman, BS[32]; Alejandra Valenzuela, BS[47]; Catherine Wandell, BS[21]; Diana Willeke[8]; Karen Williams, BA[3] and Dilinuer Wubuli, MB[43]

**Partners Scientific Advisory Board (Acknowledgement)**

**Funding: PPMI – a public-private partnership – is funded by the Michael J. Fox Foundation for Parkinson's Research and funding partners, including 4D Pharma, Abbvie, AcureX, Allergan, Amathus Therapeutics, Aligning Science Across Parkinson's, AskBio, Avid Radiopharmaceuticals, BIAL, BioArctic, Biogen, Biohaven, BioLegend, BlueRock Therapeutics, Bristol-Myers Squibb, Calico Labs, Capsida Biotherapeutics, Celgene, Cerevel Therapeutics, Coave Therapeutics, DaCapo Brainscience, Denali, Edmond J. Safra Foundation, Eli Lilly, Gain Therapeutics, GE HealthCare, Genentech, GSK, Golub Capital, Handl Therapeutics, Insitro, Jazz Pharmaceuticals, Johnson & Johnson Innovative Medicine, Lundbeck, Merck, Meso Scale Discovery, Mission Therapeutics, Neurocrine Biosciences, Neuron23, Neuropore, Pfizer, Piramal, Prevail Therapeutics, Roche, Sanofi, Servier, Sun Pharma Advanced Research Company, Takeda, Teva, UCB, Vanqua Bio, Verily, Voyager Therapeutics, the Weston Family Foundation and Yumanity Therapeutics.**

1 Institute for Neurodegenerative Disorders, New Haven, CT
2 University of Luxembourg, Luxembourg
3 Northwestern University, Chicago, IL
4 University of Iowa, Iowa City, IA
5 VectivBio AG
6 The Michael J. Fox Foundation for Parkinson's Research, New York, NY
7 Innsbruck Medical University, Innsbruck, Austria
8 Paracelsus-Elena Klinik, Kassel, Germany
9 University of California, San Francisco, CA
10 Laboratory of Neuroimaging (LONI), University of Southern California
11 BioRep, Milan, Italy
12 University of Pennsylvania, Philadelphia, PA
13 National Institute on Aging, NIH, Bethesda, MD
14 Mount Sinai Beth Israel, New York, NY
15 Indiana University, Indianapolis, IN
16 Emory University of Medicine, Atlanta, GA
17 Oregon Health and Science University, Portland, OR
18 University of Alabama at Birmingham, Birmingham, AL
19 University of South Florida, Tampa, FL
20 Baylor College of Medicine, Houston, TX
21 University of Washington, Seattle, WA
22 Boston University, Boston, MA
23 University of Rochester, Rochester, NY
24 Banner Research Institute, Sun City, AZ
25 Cleveland Clinic, Cleveland, OH
26 University of Tübingen, Tübingen, Germany
27 University of California, San Diego, CA
28 Johns Hopkins University, Baltimore, MD
29 Imperial College of London, London, UK
30 University of Salerno, Salerno, Italy
31 Parkinson's Disease and Movement Disorders Center, Boca Raton, FL
32 University of Cincinnati, Cincinnati, OH
33 Hospital Clinic of Barcelona, Barcelona, Spain
34 Hospital Universitario Donostia, San Sebastian, Spain
35 Tel Aviv Sourasky Medical Center, Tel Aviv, Israel
36 National and Kapodistrian University of Athens, Athens, Greece
37 Columbia University Irving Medical Center, New York, NY
38 Stanford University, Stanford, CA
39 University of Pittsburgh, Pittsburgh, PA
40 Center for Strategy Philanthropy at Milken Institute, Washington D.C.
41 Rutgers University, Robert Wood Johnson Medical School, New Brunswick, New Jersey
42 University of Michigan, Ann Arbor, MI
43 Toronto Western Hospital, Toronto, Canada
44 The Ottawa Hospital, Ottawa, Canada
45 Massachusetts General Hospital, Boston, MA
46 University of Kansas Medical Center, Kansas City, KS
47 University of Southern California, Los Angeles, CA
48 Barrow Neurological Institute, Phoenix, AZ

49 Mayo Clinic Arizona, Scottsdale, AZ
50 University of Colorado, Aurora, CO
51 NYU Langone Medical Center, New York, NY
52 University of Florida, Gainesville, FL
53 Montreal Neurological Institute and Hospital/McGill, Montreal, QC, Canada
54 Cleveland Clinic-Las Vegas Lou Ruvo Center for Brain Health, Las Vegas, NV
55 Clinical Ageing Research Unit, Newcastle, UK
56 John Radcliffe Hospital Oxford and Oxford University, Oxford, UK
57 Universität Lübeck, Luebeck, Germany
58 Radboud University, Nijmegen, Netherlands
59 TransThera Consulting
60 Duke University, Durham, NC
61 Wolfson Institute of Population Health, Queen Mary University of London, UK
62 Philipps-University Marburg, Germany
63 University of Lagos, Nigeria
